# Impact of Immediate and Preferential Relaxation of Social and Travel Restrictions for Vaccinated People on the Spreading Dynamics of COVID-19 : a Model-Based Analysis

**DOI:** 10.1101/2021.01.19.21250100

**Authors:** B Shayak, Mohit M Sharma, Anand K Mishra

## Abstract

**Background:** Four COVID-19 vaccine candidates developed by Pfizer, Moderna, University of Oxford/ Astra Zeneca (also Oxford/ Serum Institute of India) and ICMR/ Bharat Biotech have been granted emergency use authorization in the democratic world following established clinical trial procedures in their respective countries. Vaccination of the general public is expected to begin in several weeks. We consider the question of whether people who have received the vaccine can be selectively and immediately cleared to return to normal activities, including hassle-free travel.

**Methods:** We use a delay differential equation model developed previously by our group to calculate the effects of vaccinee “immunity passports” on the spreading trajectories of the disease. We consider default virus strains as well as high-transmissibility variants such as B1.1.7 in our analysis.

**Results:** We find that with high vaccine efficacy of 80 percent or greater, vaccinees may be immediately cleared for normal life with no significant increase in case counts. Free travel of such vaccinees between two regions should not jeopardize the infection control performance of either. At current vaccine administration rates, it may be eight months or more before COVID-19 transmission is significantly reduced or eliminated. With lower vaccine efficacy of approximately 60 percent however, social as well as travel restrictions for vaccinees may need to remain in place until transmission of the disease is eliminated.

**Conclusions:** Designing high-efficacy vaccines with easily scalable manufacturing and distribution capacity should remain on the priority list in academic as well as industrial circles. Performance of all vaccines should continue to be monitored in real time during vaccination drive with a view to analysing socio-demographic determinants if any of efficacy, and optimizing distribution accordingly. A speedy and efficacious vaccination drive will provide the smoothest path out of the pandemic with the least additional caseloads, death toll and socioeconomic cost.

## Introduction

Over the past month, four vaccine candidates have received emergency use authorization in democratic countries following rigorous trial procedures and have started being administered worldwide. These are the Pfizer and Moderna candidates based on the mRNA platform, the University of Oxford/ Astra Zeneca/ Serum Institute of India candidate based on viral vector and the ICMR/ Bharat Biotech candidate based on inactivated whole virus. The first two candidates^1,2^ have reported efficacies of almost 95 percent in phase 3 clinical trials. The third candidate^3^ has reported 60 percent efficacy with the intended dosing regimen and 90 percent with an unplanned administration regimen. More trials are underway to determine the optimal dosing pattern. The fourth candidate^4,5^ has reported encouraging results in the early trials; participant enrolment for the phase 3 trial has been completed but the trial itself has not. Thus, the efficacy data for the Oxford/ AZ/ SII and ICMR/ BB vaccine candidates are not final yet. Over and above this, Russia has approved a vector vaccine called Sputnik 5 bypassing some of the trial protocols; formal validation of this vaccine is currently underway in other countries. From the cold chain perspective, the Pfizer vaccine requires to be stored at approximately −75 °C, the Moderna candidate requires −20 °C while the Oxford/ AZ/ SII and ICMR/ BB candidates can be stored at +2 to +8 °C; Sputnik again needs −18 °C or lower.

Mathematical modeling studies of COVID-19 dynamics post-vaccination started emerging as soon as the first vaccines were approved. Swan, Goyal, Bracis et. al.^6^ have performed a detailed analysis of the roles played by different vaccine efficacy metrics. Several studies^7-10^ find that vaccinating high-contact people first will have the greatest beneficial effect on the spread of the disease. Foy, Wahl, Mehta et. al.^11^ find that priority vaccination of elderly and vulnerable people is best for reducing COVID-19 deaths. They also find that continuing with social restrictions such as six-foot separation and mask regulations during the vaccination drive will best mitigate disease spread. This latter point has been stated more emphatically by Galanti, Pei, Yamana et. al.^12^ who find almost zero difference between vaccination and no vaccination if all non-pharmaceutical interventions are relaxed. Moore, Hill, Tildesley et. al.^13^ have an equally bleak outlook for a 75 percent effective vaccine with high surge in cases and deaths if social restrictions are relaxed even one year after vaccination starts. Grundel et. al.^9^ also advocate the enforcement of social restrictions during the vaccination drive. Matrajit and Eaton^14^ find a similar conclusion applicable to long term care facilities. A more optimistic view may be found in Alvarez, Bravo-Gonzalez and Trujillo-de Santiago^15^ who recommend strong social restrictions for only the first three months of vaccination and Betti, Bragazzi, Heffernan et. al.^16^ who again permit a gradual relaxation of non-pharmaceutical interventions starting from the fourth month.

From the general public’s perspective, continued social restrictions for vaccinees appears extremely disruptive. If we have got the vaccine, we would at least hope to socialize freely with others who have been vaccinated as well. We would also hope to travel without testing for virus before and after the journey, wearing a mask in an already claustrophobic airplane economy class cabin or quarantining for days upon arrival. From an economic perspective, social restrictions during vaccination drive will amount to continued strain on the state’s fiscal resources – any relaxation or exemption will act as a lifeline. Our quest here is to find such an exemption – specifically, we ask whether social restrictions can be immediately and preferentially relaxed for those individuals who have received the vaccine. Hereafter, we refer to this strategy as **“selective relaxation”**. With ideal vaccines, the answer to this question would have been obvious. However, the actual COVID-19 vaccines are not 100 percent efficacious, which raises the issue of whether unrestricted (or at least significantly expanded) social activity and mobility on the part of vaccinees may fuel a wave of cases. We address this question in the remainder of this Article.

## Methods

We use a compartmental or lumped parameter delay differential equation model developed by our group to analyse the various situations of interest. We have selected this model because all parameters here are directly related to the disease or to control measures^17^. This feature enables the model to generate realistic epidemiological curves with default assumptions^18,19^ and make subtle predictions regarding the spreading trajectories with temporary immunity^20^. Moreover, the model is easily generalizable to new situations, with vaccination and travel being the ones considered in this Article.

We present here only the outline of the model, with the full equations and derivation being given in §1 of the Supplementary Data. The model is applicable to any region with good interaction among its inhabitants. The baseline model (with neither vaccines nor travel) features a single dependent variable *y*(*t*), the cumulative count of COVID-19 cases as a function of time. With vaccines but no travel, we require three dependent variables : *y*(*t*) the cumulative count of cases among the non-vaccinated group, *z*(*t*) the cumulative count of cases among the vaccinated group and *v*(*t*) the cumulative number of people who have been vaccinated. We classify a person as vaccinated only after s/he has received the second dose of a two-dose regimen and cleared the subsequent immunogenicity period of one or two weeks, depending on the vaccine type. To model travel, we consider two regions, City 1 and City 2, which are connected through travel links; in this case, our model features eight variables in total.

### The major parameters of interest are as follows

- Vaccine efficacy *η* : We define this as the probability that a vaccinated person is immune to the disease. Here we assume that vaccines confer either full sterilizing immunity or zero transmissibility-reducing immunity.
- Mobility index *R*_*eq*_ : Equivalent reproduction number *R*_*eq*_ is best explained through an example. The statement “vaccinees have an *R*_*eq*_ of 3” means that vaccinated people interact with others at such a rate that, were all cases to have that interaction rate and were everyone else to be susceptible, then the reproduction number *R* of the disease would have been 3. We give the mathematical definition of *R*_*eq*_ in §1 of the Supplementary Data; here we emphasize that it is a mobility index and not the actual reproduction number characterizing the outbreak.
- Travel rate *γ* : This is the fraction of all people of one region who travel to the other region each day. Since it is typically very small, we measure it in basis points i.e. one percent of a percent.
- Travel amplification factor *k* : A COVID-19 case travelling on board a flight, train or bus might spread the disease to other passengers in the vehicle. We define *k* to be the number of additional passengers who get infected by one travelling case, assuming that the former are all susceptible.

We shall solve the model using numerical integration in the software Matlab. The method will be 2^nd^ order Runge Kutta with a time step of 1/1000 day. The spatial domain of solution will be a Notional City of total population 3,00,000; we shall assume everyone to be susceptible initially and shall seed the model with a small number of initial cases. We shall run the simulations until the region has less than one active case for fourteen consecutive days, at which point we shall terminate the run and declare the epidemic to be over. Details of the initial and terminal conditions are again given in §1 of the Supplementary Data.

## Results

We separately consider the two situations where there is no travel (or equivalently, all travellers are quarantined) and where there is travel. In each case, the parameters we shall use to measure the infection control performance will be the duration *T* of the outbreak, the total case count *X* at the end of the outbreak, the vaccinated case count *Z* at the end of the outbreak and the vaccination fault ratio *f*, defined as the maximum ratio of vaccinated cases to total vaccinees at any stage in the outbreak. Since *f* is small, we report it as a percentage.

### No travel

The model we use is Equations (5-7) from the Supplementary Data. In this model, people who have not been vaccinated have a low interaction rate with other people (both vaccinees and non-vaccinees) while those who have been vaccinated have a higher interaction rate with others (again of both classes). Let the vaccines be distributed at the constant rate 600/day, which amounts to vaccination of the entire population in 500 days – a realistic estimate for many advanced countries. The vaccine efficacy shall remain variable. Let the unvaccinated people have a mobility level corresponding to *R*_*eq*_=1·15; this describes severe restrictions but not a full lockdown, since the latter can drive *R* below unity. The mobility of the vaccinated people shall again remain variable.

Table 1 presents the region’s infection control performance for three different vaccine efficacies and three values of vaccinee mobility.

**Table 1:**
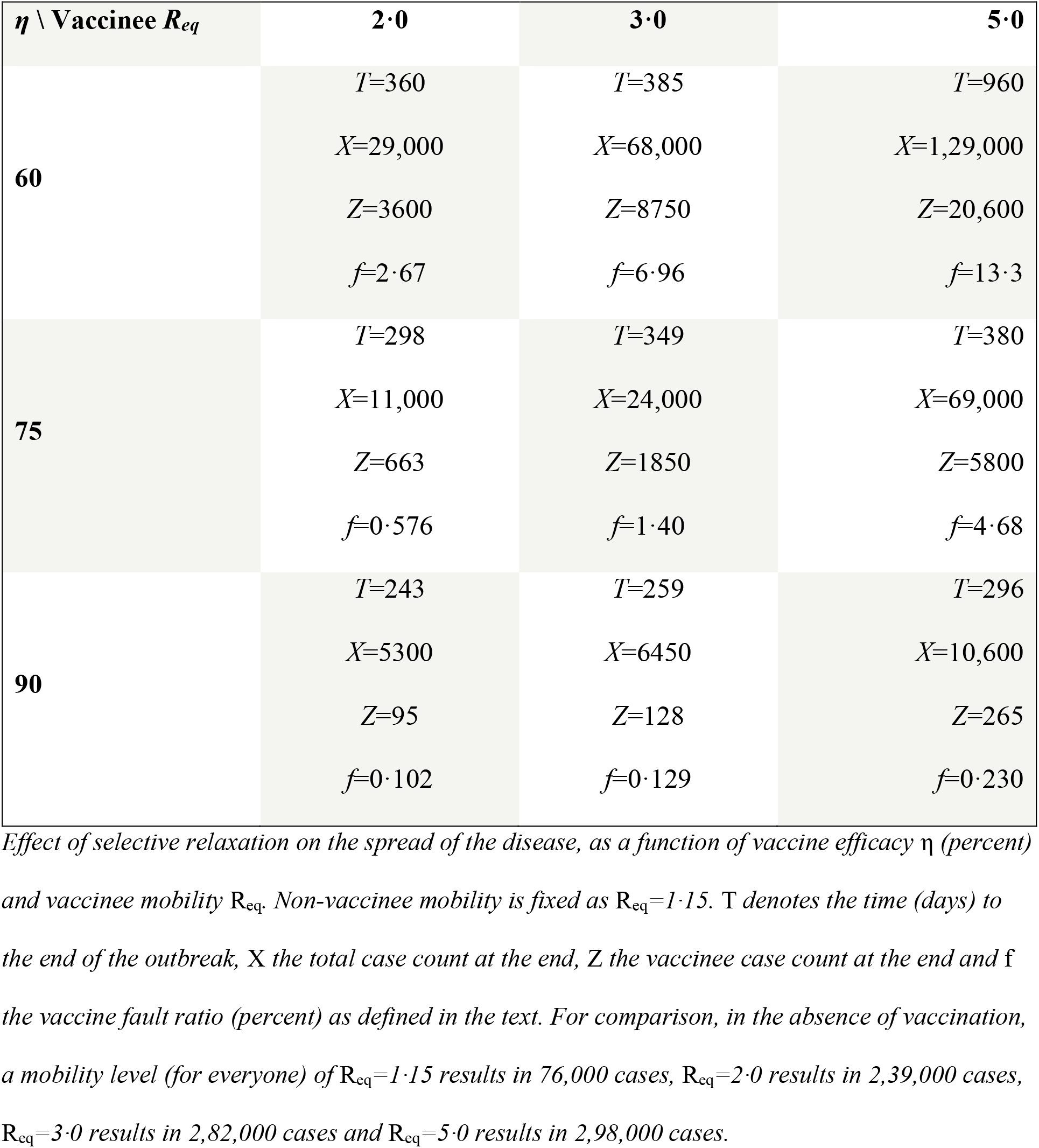
Selective relaxation performance

| $\eta \setminus \text{Vaccinee } R_{eq}$ | 2.0 | 3.0 | 5.0 |
| --- | --- | --- | --- |
| 60 | $T=360$ | $T=385$ | $T=960$ |
| | $X=29,000$ | $X=68,000$ | $X=1,29,000$ |
| | $Z=3600$ | $Z=8750$ | $Z=20,600$ |
| | $f=2.67$ | $f=6.96$ | $f=13.3$ |
| 75 | $T=298$ | $T=349$ | $T=380$ |
| | $X=11,000$ | $X=24,000$ | $X=69,000$ |
| | $Z=663$ | $Z=1850$ | $Z=5800$ |
| | $f=0.576$ | $f=1.40$ | $f=4.68$ |
| 90 | $T=243$ | $T=259$ | $T=296$ |
| | $X=5300$ | $X=6450$ | $X=10,600$ |
| | $Z=95$ | $Z=128$ | $Z=265$ |
| | $f=0.102$ | $f=0.129$ | $f=0.230$ |

*Effect of selective relaxation on the spread of the disease, as a function of vaccine efficacy $\eta$ (percent) and vaccinee mobility $R_{eq}$ . Non-vaccinee mobility is fixed as $R_{eq}=1.15$ . $T$ denotes the time (days) to the end of the outbreak, $X$ the total case count at the end, $Z$ the vaccinee case count at the end and $f$ the vaccine fault ratio (percent) as defined in the text. For comparison, in the absence of vaccination, a mobility level (for everyone) of $R_{eq}=1.15$ results in 76,000 cases, $R_{eq}=2.0$ results in 2,39,000 cases, $R_{eq}=3.0$ results in 2,82,000 cases and $R_{eq}=5.0$ results in 2,98,000 cases.*

In Figure 1, we present a typical time trace of the disease evolution.

**Figure 1:**
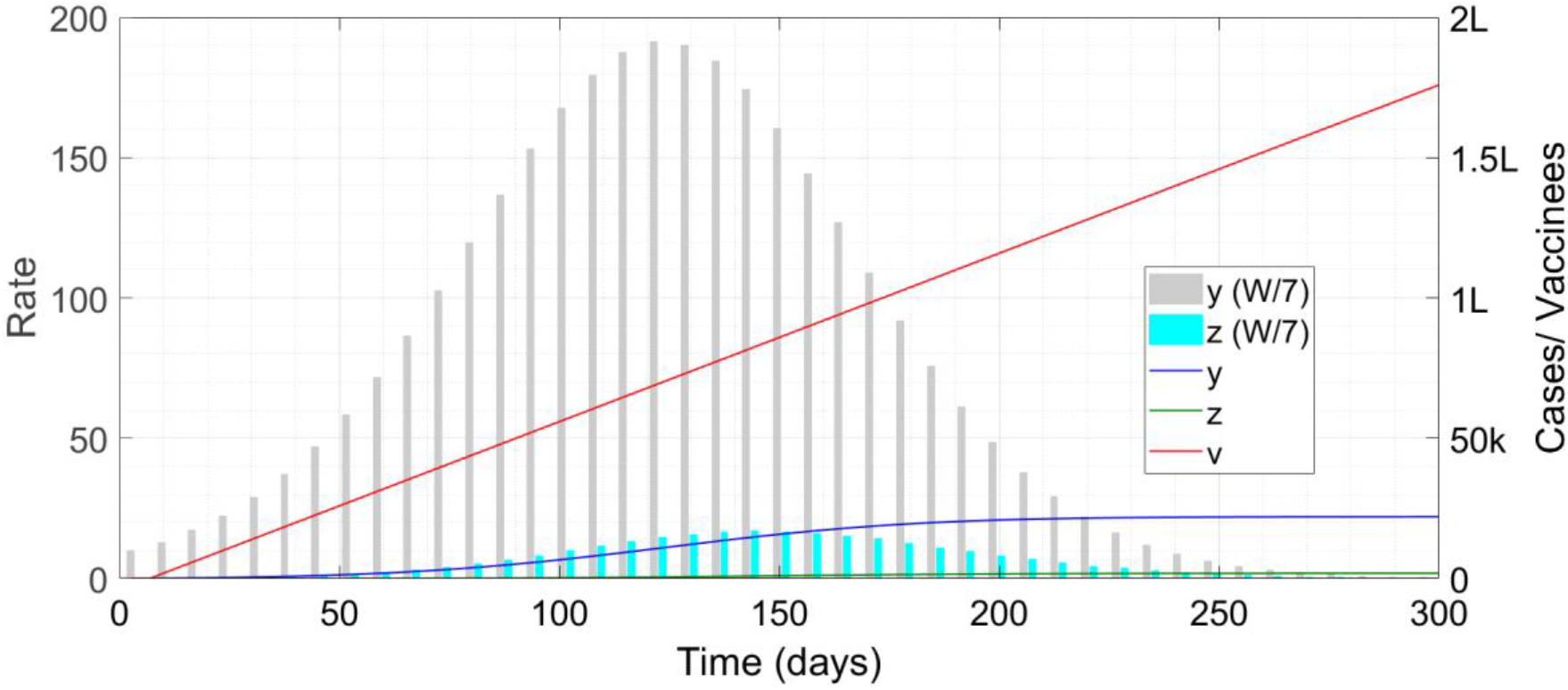
Evolution of the epidemic for the situation considered in the central cell of Table 1 i.e. η=75 percent and vaccinee mobility R_eq_=3·0. Blue line denotes unvaccinated cases, green line vaccinated cases and red line the total number of vaccinees. Grey and cyan bars denote the epidemiological curve or epi-curve, i.e. the weekly case counts in the unvaccinated and vaccinated groups respectively. We have scaled them down by a factor of seven so that the envelopes of the bars become the derivatives 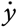 and ż. The legend W/7 indicates this. ‘k’ denotes thousand and ‘L’ hundred thousand.

We can see that although the outbreak technically lasts for 352 days, the epi-curve has become of negligible size by the 300-day mark.

### Travel

We now consider the case where two cities with potentially different infection control performance are linked by travel. As mentioned in the Methods Section, the key parameters characterizing travel are the rate *γ* and the amplification factor *k*. Since we want to examine whether vaccinees can travel without hassle, we shall consider only the case where vaccinated people travel between the two cities. The model is Equation (19) from the Supplementary Data.

We shall consider situations where two cities taken from Table 1 are coupled. In each case, City 1 shall be the better performer and City 2 the worse performer. Our primary objective will be to find whether the ingress of travellers from City 2 can spoil the infection control performance of City 1. Again, we shall present the data in a tabular form, with *γ* and *k* being the variable parameters; in each cell we shall report the end-stage infection control performance metrics for City 1. For the first set of runs, Table 2, we let *R*_*eq*_=3·0 in both cities; let City 1 have 90 percent vaccine efficacy and City 2 have 75 percent.

**Table 2:** Effects of travel 1

| $\gamma \backslash k$ | 2 | 5 | 10 |
| --- | --- | --- | --- |
| **20** | $T=347$ | $T=347$ | $T=347$ |
| | $X=6800$ | $X=7000$ | $X=7300$ |
| | $Z=142$ | $Z=151$ | $Z=165$ |
| | $f=0.134$ | $f=0.138$ | $f=0.147$ |
| **50** | $T=345$ | $T=345$ | $T=345$ |
| | $X=7300$ | $X=7800$ | $X=8600$ |
| | $Z=164$ | $Z=185$ | $Z=220$ |
| | $f=0.143$ | $f=0.154$ | $f=0.173$ |
| **100** | $T=342$ | $T=343$ | $T=345$ |
| | $X=8000$ | $X=9000$ | $X=10,700$ |
| | $Z=199$ | $Z=240$ | $Z=309$ |
| | $f=0.158$ | $f=0.182$ | $f=0.226$ |

*Infection control performance of City 1 ( $\eta=90$ percent, $R_{eq}=3$ ) as a result of travel coupling with City 2 ( $\eta=75$ percent, $R_{eq}=3$ ), as a function of the travel rate $\gamma$ (basis points) and the amplification factor $k$ . The metric $T$ denotes the time (days) to the end of the outbreak, $X$ the total case count at the end, $Z$ the vaccinee case count at the end and $f$ the vaccine fault ratio (percent), all in City 1. For comparison, in the absence of travel, City 1 has total 6450 cases over 259 days while City 2 has total 24,000 cases over 349 days.*

In Table 3, we consider a very similar scenario except that we reduce the vaccine efficacy in City 2 to 60 percent.

**Table 3:** Effects of travel 2

| $\gamma \backslash k$ | 2 | 5 | 10 |
| --- | --- | --- | --- |
| **20** | $T=384$ | $T=384$ | $T=384$ |
| | $X=8300$ | $X=9500$ | $X=11,000$ |
| | $Z=202$ | $Z=254$ | $Z=336$ |
| | $f=0.165$ | $f=0.200$ | $f=0.258$ |
| **50** | $T=382$ | $T=382$ | $T=383$ |
| | $X=11,000$ | $X=14,000$ | $X=18,000$ |
| | $Z=305$ | $Z=423$ | $Z=602$ |
| | $f=0.232$ | $f=0.316$ | $f=0.447$ |
| **100** | $T=379$ | $T=380$ | $T=383$ |
| | $X=14,000$ | $X=19,000$ | $X=27,000$ |
| | $Z=463$ | $Z=673$ | $Z=980$ |
| | $f=0.336$ | $f=0.484$ | $f=0.705$ |

*Infection control performance of City 1 ( $\eta=90$ percent, $R_{eq}=3$ ) as a result of travel coupling with City 2 ( $\eta=60$ percent, $R_{eq}=3$ ), as a function of the travel rate $\gamma$ (basis points) and the amplification factor $k$ . The metric $T$ denotes the time (days) to the end of the outbreak, $X$ the total case count at the end, $Z$ the vaccinee case count at the end and $f$ the vaccine fault ratio (percent), all in City 1. For comparison, in the absence of travel, City 1 has total 6450 cases over 259 days while City 2 has total 68,000 cases over 385 days.*

This completes the presentation of the most salient results – for a variety of other results and their interpretation we must refer to §3 of the Supplementary Data.

## Discussion

We again consider separately the cases without and with travel.

### No travel

Analysis of the initial spread of COVID-19 before any kind of restrictions were applied^21-24^ has yielded a reproduction number of approximately 3. Hence the situation *R*_*eq*_=3, which we have considered in Table 1, is indicative of completely normal activity on the part of the vaccinees, with the original strain/s of the virus. The situation *R*_*eq*_=5 might correspond to normal activity with a highly transmissible virus variant such as the B1.1.7^25^ or N501Y^26^ form, while *R*_*eq*_=2 can indicate either normal life with a low-transmissibility strain or partial social restrictions with the other strains.

We can see that with selective relaxation, the 60 percent effective vaccine causes less than 10 percent final infection level only for the lowest of the three *R*_*eq*_’s considered while the 90 percent effective vaccine results in a very low infection count in all cases. With a highly effective vaccine, selective relaxation appears to be a safe and robust reopening strategy which leads to elimination of the disease in time while constantly expanding socioeconomic activities. A general lifting of restrictions can be announced when disease transmission has significantly reduced or ceased. We can qualitatively explain the effectiveness of the selective relaxation strategy with the help of two arguments.

The first argument focusses on the reproduction number *R*. With no interventions, COVID-19 has an *R*_0_ of 3 means that, when everyone is susceptible, one person spreads the disease to three other people during the course of everyday interaction. When vaccinees interact normally among each other, if two out of every three are immune (i.e. vaccine efficacy is 67 percent) – or more – then *R* will decrease below unity and the epidemic will die out in time.

The second argument focusses on an individual vaccinee, whom we call A. Vaccine efficacy of 90 percent implies 10 percent failure probability which does not sound very small. However, 10 percent is the probability that A catches the disease **given** an exposure. Transmission is a two-person process – if A interacts only with other vaccinees, then the probability that they have the disease and can expose A to the pathogen also reduces to (approximately) 10 percent. A’s total contraction probability therefore reduces to approximately 1 percent i.e. the disease contraction probability is quadratic and not linear in the vaccine failure probability.

Of course, during any reopening activity, the case counts in both unvaccinated and vaccinated populations will need to be monitored continuously. Given a time series, our model can be used to predict the future performance, and restrictions on vaccinees will have to be reimposed if it turns out that they are driving spread. The duration of about 250-300 days to the end of the outbreak which we have found in Table 1 is, we believe, a reasonably robust prediction. This is because a reopening plan is built into the outbreak’s evolution – it ends with society in normal life and not in a restricted mode of operation. As Table 1 shows, the duration depends strongly on the vaccine efficacy and less sensitively on the mobility of the vaccinees – further numerical work (not shown here) yields the dependence of the duration on the non-vaccinee mobility to be similarly weak. Note also that the outbreak ends at approximately 50 percent vaccination coverage, so the termination is not a “herd immunity” effect.

### Travel

In §3 of the Supplementary Data, we argue that *k*=5 is perhaps a typical situation and *k*=10 a worst-case. Similarly, 100 basis points or one percent of a city’s population travelling every day also appears like an overestimate. Hence at least one scenario of Tables 2 and 3 represents a game against nature.

In these Tables, we can see a drastic difference in City 1’s infection control performance depending on whether the vaccine efficacy in City 2 is 75 or 60 percent. In the former situation, the infection counts in all cases remain within twice of that in the absence of travel; in the latter situation, the counts may increase by as much as five times. Needless to say, travel coupling of two cities with 90 percent vaccine efficacy poses negligible risk in all cases. We can find similar results (not shown) for travel coupling between other city pairs in Table 1. In a nutshell, we can say that free travel between regions which are individually in control of their outbreaks is permissible; free travel to and from a region with an out-of-control outbreak is not. Again, monitoring of the situation will be required if travel links are opened with the disease still prevalent. We must remember that all the results are predicated on the hassle-free travel of vaccinees alone and not of a mixed group. Hence, arrangements should be made so that vaccinees do not mix with non-vaccinees during travel. For example, there can be special flights and trains for people of each category, with the vaccination status being verified together with the travelling ticket.

### Limitations

The limitations of the analysis come from the various assumptions in the model. One set of limitations is common to any lumped-parameter or compartmental model. This is that when the absolute number of cases becomes very low, the model ceases to remain valid – the deterministic evolution is replaced by a stochastic process. Hence, predictions regarding the end-stage of the disease might not be accurate. Other limitations arise from assumptions regarding vaccine immunity, vaccinee mobility etc. We have tried wherever possible to ensure that errors are on the side of caution, and have presented a detailed discussion of this aspect in §2 of the Supplementary Data.

## Conclusion

In this Article, we have identified immediate and preferential relaxation of restrictions for vaccinees as a feasible path to the elimination of the dreadful pandemic called COVID-19. This path features a continuous growth of economic and social activities during the vaccination drive. The incentive of immediate benefits will also induce people to receive the immunizations and hence automatically combat vaccine hesitancy^27^. With this mode of operation, and with current encouraging vaccine efficacies, we find a timeframe of approximately eight months before transmission reduces to negligible levels. We also find that vaccinated people can be allowed to travel freely between regions of good infection control performance. This finding should act as a boost to the travel industry which is currently reeling from the effects of severely reduced demand.

While our primary finding and its associated message is hopeful, there are also some cautionary takeaways. In particular, a 60 percent effective vaccine does not appear to be adequate for issuing immunity passports. Until and unless high-efficacy vaccines are widespread, research on improving vaccine efficacy should be pursued at full throttle. “In the field” efficacy estimations should continue for all approved vaccines, especially to identify socio-demographic determinants of efficacy if any.

In conclusion, the initiation of vaccination drives marks the beginning of the end of humanity’s struggle against COVID-19. Our immediate objective over the remaining few months of this battle has to be to minimize the caseloads, death tolls and disease burden from the socioeconomic perspective. We hope that the prescription we have suggested here may prove effective in this respect.

## Data Availability

There is NO data referred to in the manuscript

## Author contribution statement

All of us have contributed equally to the manuscript.

## SUPPLEMENTARY DATA

In this Supplement we cover several issues which could not be treated in the Article proper due to lack of space. Throughout, a figure or table numbered “n” always refers to the Article proper while a figure or table numbered “Sn” refers to this document. The same holds for References. Since equations exist in the Supplement alone, we have labelled them with numbers only and no “S” prefix.

### §1 MODEL DERIVATION AND EQUATIONS

We begin with a very brief recap of the model proposed in our prior works [17-20] (cited in the Article proper).

#### RECAPITULATION

Like every lumped-parameter or compartmental model, ours is applicable in any region with good connection among its inhabitants, such as a neighbourhood, town, village, or smaller city. Metropolitan cities may need to be partitioned into several regions, depending on internal connectivity. The model treats the transmission of disease as a process of interaction between at large cases and susceptible targets; its underlying philosophy is

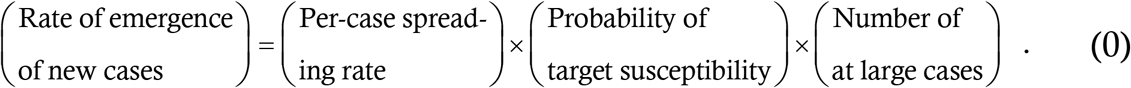

Defining *y* (*t*) as the cumulative case count, the left hand side above is d*y*/d*t*. The per-case spreading rate, which we call *m*_0_, is the product of two quantities – the rate at which a random person (and hence an at large case who is unaware of infectious nature) interacts with other people, and the probability that an interaction with a susceptible target results in a transmission. The interaction rate is governed by the degree of social restrictions in place while the transmission probability is determined by masking and sanitization; collectively, *m*_0_ embodies the effects of non-pharmaceutical interventions [17]. The target susceptibility probability factors in the immune response to the disease; with permanent immunity (and the approximations of instantaneous recovery and zero mortality), it takes the form 1 − *y*/*N* where *N* is the region’s total population. The number of at large cases has the following mathematical form :

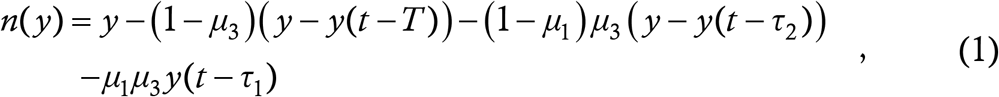

where *μ*_1_ is the fraction of cases who are asymptomatic, *μ*_3_ is the fraction of cases who escape from contact tracing, *T* is the time for which contact traced cases remain at large, *τ*_1_ is the time for which untraced asymptomatic cases remain transmissible and *τ*_2_ is the time for which untraced symptomatic cases remain transmissible before manifesting symptoms and (at least we assume) seeking quarantine.

Putting all this together, we arrive at the retarded logistic equation

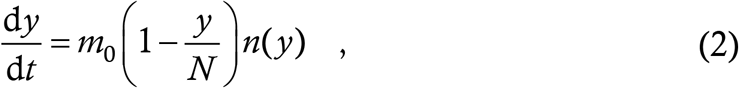

as the final form of the dynamic epidemic model in the absence of vaccination and travel. Equation (2) uses delays rather than inverse-rates to express infection durations, which enables it to make very realistic predictions. For further details of derivation, we must refer to our prior study [17].

#### VACCINATION ONLY

Here we add vaccination without travel to the basic model (2). For this, we define three dependent variables : *y* (*t*) the cumulative count of corona cases among unvaccinated people, *z* (*t*) the cumulative count of cases among vaccinated people and *v*(*t*) the total number of vaccinated people. We now use the structure (0) to formulate the evolution equations for the disease; for conceptual clarity, we permute the terms on the right hand side as follows :

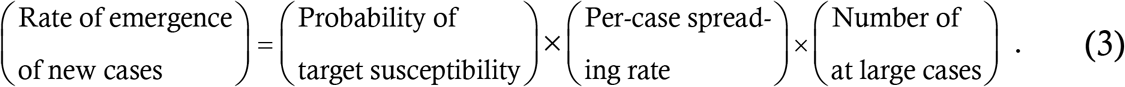

In this layout, the two terms involving cases rather than targets are adjacent to each other.

Let us now formulate the equation for *y* (*t*), the unvaccinated cases. At any time, the total number of vaccinees is *v* and the total number of non-vaccinees is *N* − *v*. In this work, we assume that the disease as well as the vaccine confers permanent immunity (the disease with 100 percent probability and the vaccine only in those instances where it works), an assumption discussed in detail in §2. An unvaccinated person can be insusceptible only if s/he has already contracted and recovered from the disease; at any time the total number of recoveries (modulo the approximations of the previous Subsection) is *y*, the total number of non-vaccinees is *N* − *v* so the probability that a random non-vaccinee is susceptible is (*N*−*v*−*y*)/(*N*−*v*), which is 1 − *y*/(*N*−*v*).

As per the model assumptions, non-vaccinees and vaccinees have different interaction rates, and hence spreading rates, with the latter being higher. Consequently we shall replace the single value *m*_0_ with two values *m*_*l*_ (low) and *m*_*h*_ (high), which will be applicable to the two categories respectively. We assume that every at large case’s *m* (either *l* or *h*) spreading incidents per day are distributed among non-vaccinees and vaccinees in proportion to their population i.e. a fraction (*N*−*v*)/*N* of any case’s transmissions are to non-vaccinees and a fraction *v*/*N* to vaccinees. We shall return to this point in §3.

As for the number of at large cases, we take the asymptomatic fraction *μ*_1_ = 4/5, which is towards the higher end of the spectrum. We take the contact traced fraction to be zero so that *μ*_3_ = 1. In Ref. [17] we have shown that contact tracing will indeed capture only a small percentage of total cases if the asymptomatic fraction is high; moreover, contact tracing is managed by healthcare professionals many of whom will now be re-deployed to vaccination drive. We use the parameter values [S1] *τ*_1_ = 7 and *τ*_2_ = 3, so that the function *n* gets defined as

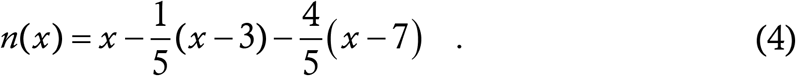

We assume that vaccinated cases have the same *μ*_1_, *τ*_2_ and *τ*_1_ as non-vaccinated ones (details in §2), so that we can use this function *n* to count at large cases of both unvaccinated and vaccinated groups. Putting all this together, we have the equation

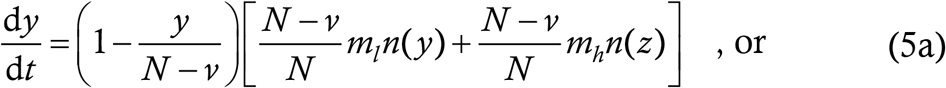

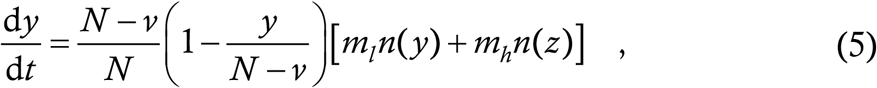

with the second line being a simple rearrangement of the first.

Similarly we can formulate the equation for the vaccinees. By the model assumptions, the vaccine confers sterilizing immunity with probability *η*, so at any time, the number of insusceptible vaccinees is *ηv* and the number of susceptible vaccinees is (1−*η*)*v*. Among the latter, *z* people have contracted and recovered from the infection so they are insusceptible as well. Hence, the total number of susceptible vaccinees is (1−*η*)*v* − *z* and the susceptibility probability is this divided by *v*, which is 1−*η* − *z*/*v*. The fraction (*N*−*v*)/*N* in (5) will get replaced by *v*/*N* and the number of at large cases will remain the same as in (5), giving

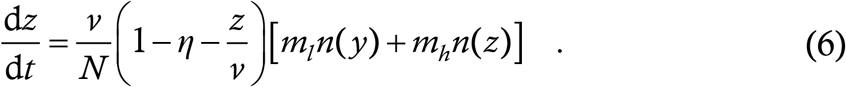

Finally, we need an equation for *v*. We assume that vaccination takes place at a constant rate *α* (people/day). The longest that the vaccination drive can continue is until all non-vaccinees have either turned into cases or vaccinees i.e. when *y* + *v* equals the total population. Thus, we have

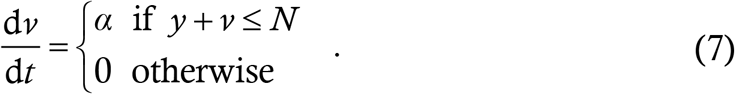

The stopping condition assumes that everyone is willing and able to receive the vaccine. It ignores ineligible people (such as children) and vaccine hesitant people. However, in almost all the runs considered in the Article, the outbreak and hence the vaccination drive are over long before the stopping condition (7) is actually attained, so this does not pose any trouble.

Equations (5-7) provide the basis for all the simulations in the “Results → No travel” Section of the Article proper. A delay differential equation needs to be seeded with an initial function having the duration equal to the maximum delay involved in the problem, which is 7 days in this instance. The initial function we have chosen is *y* (*t*) = 10*t, z* (*t*) = 0 and *v* (*t*) = 70 for *t* belongs to the interval [0, 7], with the equations themselves being solved for *t* > 7. For the termination condition, we define the active case count at time *t* to be

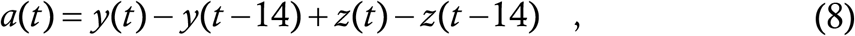

and terminate the run if *a*(*t*) < 1 for 14 consecutive days.

We now present the mathematical definition of the mobility index *R*_*eq*_. In the Article proper we have explained it through an intuitive example; here we make it rigorous. As we can see, *R*_*eq*_ does not appear as a parameter in (5-7) even though we have treated it like one in the Article. Actually, *R*_*eq*_ is directly proportional to *m* (either *l* or *h*). It can be shown [17] for the baseline model (2) that the reproduction number *R* is given by

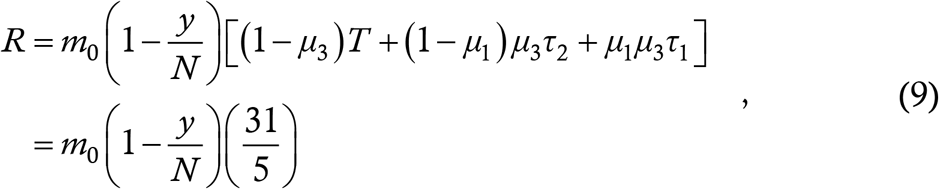

if we substitute the numerical values we are using here. The starting value *R*_0_ is defined at the beginning of the epidemic i.e. when *y*/*N* is nearly zero; thus we can say *R*_0_ = (31/5)*m*_0_. We use this proportionality to define *R*_*eq*_ in terms of *m* :

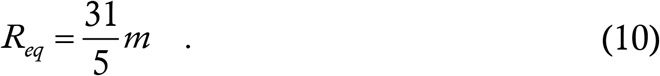

Thus, unvaccinated people have *R*_*eq*_ = 1·15 means that *m*_*l*_ = 1·15 × (5/31) = 0·186; vaccinated people have *R*_*eq*_ = 3 means that *m*_*h*_ = 3 × (5/31) = 0·484. Similarly, vaccinee *R*_*eq*_’s of 2 and 5 correspond to *m*_*h*_ values of 0·323 and 0·807 respectively.

We reiterate that *R*_*eq*_ is only an index which measures people’s mobility and is NOT the actual reproduction number of the disease during the epidemic evolution – we have expressed *m* in terms of *R* only because *m* is very difficult to measure but *R* for COVID-19 has been measured and found to be approximately 3 in the absence of social restrictions [21-24] (cited in the Article proper).

#### TRAVEL ONLY

Now we consider the case where there is travel but no vaccination. This situation will help us to develop the concepts needed to formulate the model with both vaccination and travel. We consider two regions, City 1 and City 2, which are connected by travel. We assume that the total populations of the two cities do not change as a result of travel. We shall use subscripts 1 and 2 to refer to variables related to the two cities. Let us focus on City 1 during the derivation. There are three mechanisms through which corona cases can be generated in City 1. The first is the familiar process, through internal spread of the virus in City 1. The second is through influx of cases from City 2 – since we are interested in a situation with no traveller quarantine, the imported cases will also spread virus in City 1. Indeed, at the very start of the pandemic, the outbreak in every country save one began with imported cases. The third mechanism is ancillary to the second but has a difference which we shall explain shortly. On the train or flight from City 2 to City 1, each travelling case will transmit the disease to some other people. In the Article proper, we have defined this number to be the travel amplification factor *k*. Thus, each case imported into City 1 will carry with him/her a baggage of *k* ancillary cases who will also spread the virus in City 1.

The difference between the imported and the ancillary cases is the duration for which they remain at large. Some of the imported cases will have made their journey early on in their transmissibility period – they will do the bulk of the spreading in City 1 itself. Others however will have done the bulk transmission in City 2 before boarding the inbound vehicle. On average, travelling cases will remain at large for half the total time in each city. Thus, a travelling asymptomatic case will remain at large in City 1 for 7/2 days while a travelling symptomatic case will remain at large in City 1 for 3/2 days. An ancillary case however will spend his/her entire transmissibility duration in City 1 (we assume that the duration of the journey itself is negligible compared to the transmissibility period and that a case does not travel more than once within the transmissibility period).

To mathematically account for the three generation mechanisms, we define three classes of cases in City 1, as follows :

- *y*_1_, a case who contracts the virus in City 1
- *w*_1_, a case who contracts the virus in City 2 and imports it into City 1
- *x*_1_, a case who contracts the virus from an imported case on board the inbound vehicle to City 1

As usual, the variables will stand for the cumulative counts of all the case classes. We also need a new function *ñ*(·) to describe the half-length stays in each city made by the travelling cases; analogous to (4) we define

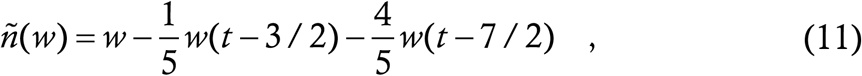

as the number of at large half-time cases in either city. We can now embark on the development of the model equations.

We start from the equation for *y*_1_. Cases generation is described as usual by (0). A question immediately arises regarding the susceptibility probability – how many of the travelled cases should we include among the immune population ? In other words, should we treat only *y*_1_ as insusceptible or *y*_1_+*w*_1_+*x*_1_ combined as insusceptible ? Also, what about the cases *y*_1_ who are lost to travel ? While it is unrealistic for a person to travel twice or more during his/her transmissibility period, it is certainly reasonable to travel multiple times during the immunity period. In the absence of detailed knowledge regarding travel patterns, we must make an approximation here. If we go with just *y*_1_, the model will likely yield more cumulative cases than reality and if we go with *y*_1_+*w*_1_+*x*_1_ then the model will yield less cases than reality. Erring on the side of caution is always preferable so we treat the susceptibility probability as 1 − *y*_1_/*N*. Fortunately, in actual runs, *w* and *x* turn out to be much smaller than *y*, so the error involved here is small.

Calculating the number of at large cases involves a further subtlety. Recall that the definition of *y*_1_ includes every case who is generated in City 1 – both the ones who stay there throughout and the ones who get exported to City 2. The former spend the whole transmissibility period in City 1 while the latter spend only half (on average), so the two categories must be considered separately. Since *w*_2_ identifies the exports from City 1, *y*_1_ − *w*_2_ must identify the others i.e. the ones who remain in City 1 throughout and transmit for the full duration. Since *y*_1_ − *w*_2_ is cumulative, the number of transmissible among these will be given by *n* (*y*_1_ − *w*_2_). The export cases *w*_2_ remain transmissible and at large for half the time, so their numbers in City 1 will be given by *ñ* rather than *n*. The import cases *w*_1_ will again spend half the duration in City 1, while ancillaries *x*_1_ will spend the full duration. Putting all this together, we get

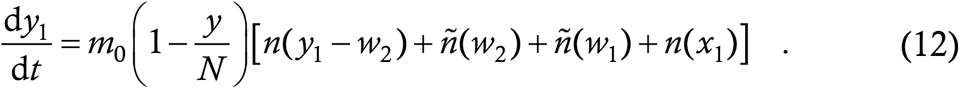

Next in the line is *w*_1_ – the cumulative count of imports from City 2. For this, we introduce the travel fraction *γ*, already defined in the Article proper as the fraction of people of each city (here 2) who travel to the other city (here 1) every day. The same fraction *γ* will apply to the corona cases unknowingly at large in City 2. Since, by our assumptions only locally generated cases travel, the rate of influx from City 2 will be *γ* times the number of at large local cases in City 2 i.e.

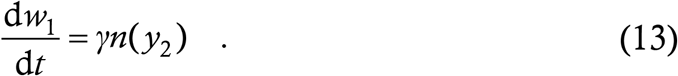

The last case class to account for is *x*. For every one imported case *w*_1_, there are *k* vehicle-spawned cases *x*_1_. Hence,

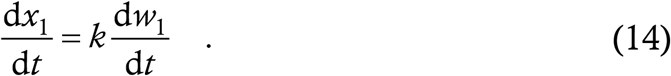

If we now assume (very plausibly) that *x*_1_(0) = *w*_1_(0) = 0 then we have a simple proportionality relation between *x*_1_ and *w*_1_. Hence we can remove *x*_1_ from the system altogether and replace it with *kw*_1_ wherever it appears. That is, we can rewrite (12) as

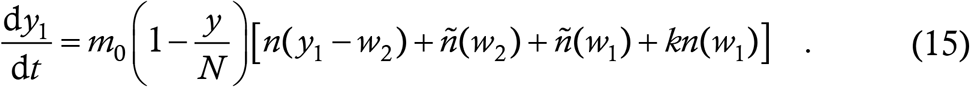

With this, the equations for City 1 are complete.

By analogy, we can write for City 2

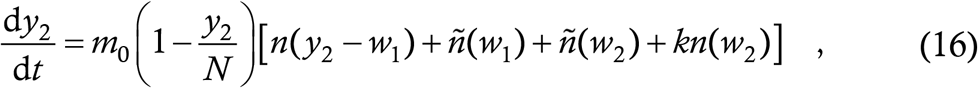

and

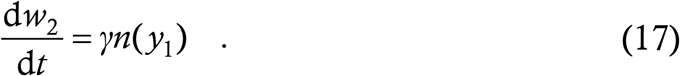

Equations (13,15-17) constitute a coupled fourth-order system describing the spreading dynamics of coronavirus in two regions connected by travel. Here we have assumed that *N* and *γ* are the same for both cities; if that is not the case then we must ensure *γ*_1_*N*_1_ = *γ*_2_*N*_2_ so that the numbers of people travelling both ways are equal.

#### VACCINATION AND TRAVEL

We now combine the two preceding derivations to formulate the model for this situation. Fortunately, almost all the work has already been done. In each city we now need five compartments – *y* : locally generated unvaccinated case, *z* : locally generated vaccinated case, *w* : imported case, *x* : ancillary travel-generated case, *v* : vaccinee. Once again, *x* becomes proportional to *w* through the constant *k* (or rather *k*’, see below) and the variable count per city reduces to 4.

The intention of our analysis is to study the effects of hassle-free travel of vaccinees, so we can ignore non-vaccinee travel altogether. We shall again need two spreading rates *m*_*l*_ and *m*_*h*_ for unvaccinated and vaccinated people. Since all travellers are vaccinated, they will have the high spreading rate in both source and destination cities. In the susceptibility probability, we will again include only the locally generated cases and none of the travelled cases. The definition of the amplification factor *k* assumes that every target is susceptible; this must be modified to account for the vaccine. If both cities have the same vaccine efficacy *η*, then *k* must get replaced by *k* (1−*η*); if the cities have different efficacies *η*_1_ and *η*_2_ then we will reduce *k* by the average efficacy i.e. replace *k* with *k*’ where

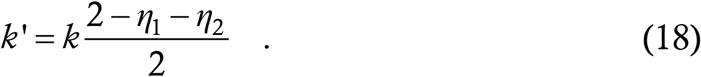

This assumes (realistically) that half of the passengers on the vehicle have one efficacy and half have the other efficacy.

We can now present the model. It is

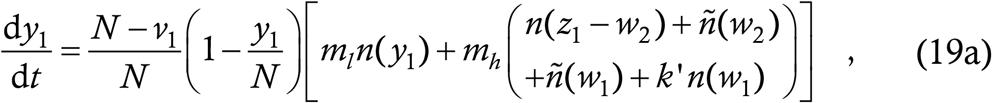

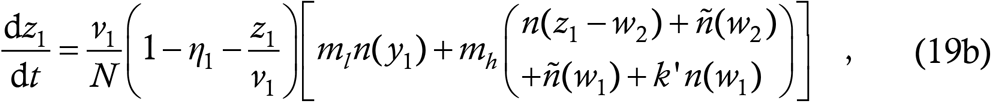

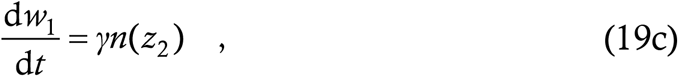

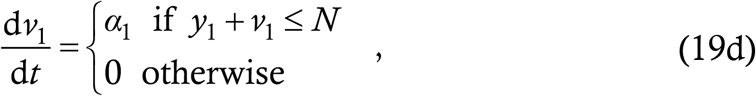

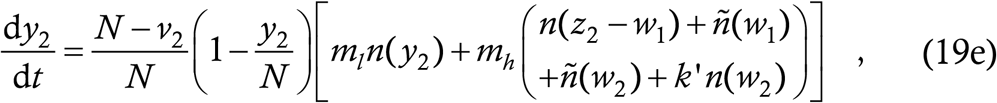

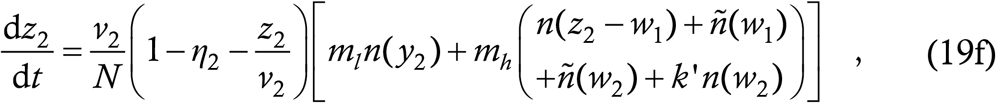

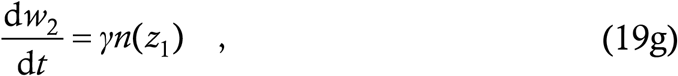

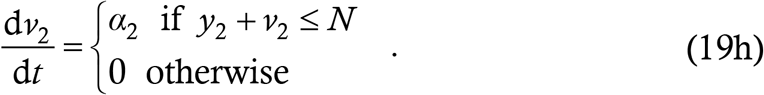

All simulation runs presented in the “Results → Travel” Section of the Article proper are based on the above eight coupled equations.

For each simulation, we have taken the initial conditions in the interval *t* belongs to [0, 7] as *y*_1_ = *y*_2_ = 10*t, v*_1_ = *v*_2_ = 70 and *z*_1_ = *w*_1_ = *z*_2_ = *w*_2_ = 0, and have solved (19) for *t* > 7. For the termination condition, we again define active cases as

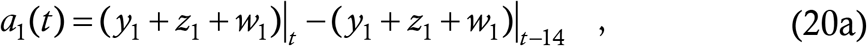

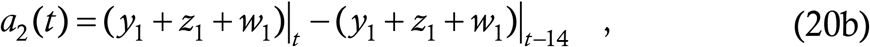

and stop the run if both *a*_1_ and *a*_2_ are less than unity for 14 consecutive days.

### §2 MODEL ASSUMPTIONS AND THEIR EFFECTS

Here we discuss some of the assumptions and approximations inherent in the model, the procedures for relaxing them, and the effects which they have on the results. For assumptions built into in the baseline model (2), we refer to Refs. [17,18]; here we consider only the approximations involved in extending the baseline to the vaccination and travel cases.

#### Vaccine immunity

As mentioned in §1, we have assumed that vaccine confers sterilizing immunity, i.e. complete immunity against contraction, symptomaticity and transmission of COVID-19, with probability *η* and zero immunity against contraction and transmission with probability 1−*η*. Currently, the Pfizer and Moderna trials [1,2] have focussed only on reduction of symptomatic infections. The Oxford vaccine [3] has included asymptomatic cases in the phase 3 analysis and found only a 30 percent reduction in asymptomatic infections in the vaccine group relative to the placebo group (compared to a 60 percent reduction in symptomatic infections). However, the absolute number of asymptomatic cases found in this study is quite small, so more data needs to be collected on this issue.

If the Oxford results are indicative of a general trend, then our assumption of equal immunity across all case classes will result in an undercounting of cases since our model will yield fewer asymptomatic vaccinated cases than reality. This is counterbalanced by our assumption that vaccinated cases have the same transmission properties (transmissibility and duration) as unvaccinated ones. It is possible (and intuitive) that vaccine cases will actually have lower viral loads and faster recovery period, which generate an error in the opposite direction. To the best of our knowledge, so far, there is nowhere near enough information which can enable us to determine more appropriate parameter choices. However, when such information does become available, it will not be difficult to incorporate it into the model by changing the values of *η* and of *μ*_1_, *τ*_1_ and *τ*_2_ for vaccinated cases.

In our model we have not made any comment regarding the severity-reducing immunity of the vaccine. That would have been relevant for counting hospitalizations and deaths among the vaccine group, which we have not attempted. In the runs we have performed however, this quantity is of secondary importance. For in Table 1 with 75 and 90 percent efficacy vaccine, we see that an overwhelming majority of the total cases occurs in the unvaccinated group, so the bulk hospitalizations and deaths will also come from that group. Only with the 60 percent effective vaccine do we see an appreciable count of vaccinated cases – even then, they are an absolute minority. It remains true however, that if the vaccine has a certain or near-certain rate of reducing hospitalizations and deaths (as seems to be the case with all the vaccines released so far), then that will generate considerable mental peace during the selective relaxation process and will enable entities like universities where many/all have been vaccinated to reopen without a second’s thought.

A further assumption we have made is that the immunity conferred by the disease as well as the vaccine is permanent. This assumption is valid so long as the immunity duration is longer than the evolution time of the outbreak, which is eight months to a year in the situations we have considered. For the disease itself, antibodies as well as cellular immune responses do seem to be durable over at least a 6-7 month period, the longest studied so far (a mini-review of literature on this topic appears in Ref. [20], while Ref. [S2] is a recent update). As for the vaccine, Moderna [S3] and ICMR/BB [5] have reported durable immune responses for at least 3 months, with the titre profiles being similar to those generated by symptomatic COVID-19 infection. Time alone will tell us the durability of vaccine immune response, but so far we see no reason to deviate from the permanent immunity assumption. We hope that it shall prove possible to accelerate the vaccination drive with time and ensure that the disease is eliminated before immunity runs out.

#### Initial and terminal conditions

The assumption that there are zero pre-existing cases at the start of the vaccination drive is an under-estimate; in some regions at least, a significant fraction of the population has already been immunized. In other regions however, the immunized fraction might not be too large. Pre-existing recoveries can influence the case trajectories in two ways : (*a*) for given *m*_*l*_ and *m*_*h*_ it can make the actual reproduction number lower than the model and hence terminate the epidemic faster and with lower caseload, (*b*) it can achieve the reproduction numbers of our simulations at higher levels of mobility especially among non-vaccinees and hence equal our infection control performance at a lower level of intervention. At the same time however, the presence of a significant number of at large cases when the vaccination drive starts can increase the vaccination fault ratio beyond the model predictions. The assumption of 70 pre-existing vaccinees has no impact other than to prevent division by zero when calculating the fault ratio.

The terminal condition of less than one active case for a sufficiently long time is an eminently plausible measure of the true end of the outbreak. The number 14 (twice) in the definition of the condition might appear somewhat arbitrary. The choice is harmless since changing that number changes the cumulative case counts by minuscule amounts. At any rate, when the absolute number of cases is very low, a lumped parameter model breaks down. All that one can talk about are probabilities, and for that one needs an agent-based model. Our model (and **any** other differential equation model) is good only for predicting when transmission will have become significantly reduced, and for that any physically plausible termination condition is adequate.

##### Vaccination fault ratio

The question we want to address is “If I receive the vaccine and move about freely, what is the probability that I shall actually contract the disease during the evolution of the outbreak ?” As discussed in the main Article, the complement of the efficacy i.e. 1−*η* is a gross overestimate since it does not factor in the two-person nature of transmission. There is no single metric in fact which can help us to answer this question. An approximate indicator will be the ratio of the total number of vaccinated cases to the total number of vaccinees. However, this index will be artificially lowered by the fact that during the tail-phase of the epidemic, there are hardly any new cases but lots of new vaccinees. Hence we have opted to evaluate the ratio at every point during the disease evolution and report its maximum value as the vaccination fault. Other metrics can also be calculated from data sets, if desired. In the “Results → Travel” section of the main Article, we have calculated the fault *f* as the maximum of the ratio *z*_1_/*v*_1_, ignoring the imported and travel-generated cases. Any error arising from this will be small and will not change the fact that the fault is always 1 percent or less.

##### Vulnerability- or transmissibility-structuring

The effects of COVID-19 on people of different age groups are widely heterogeneous. Similarly, there are some people whose profession forces them to interact with many others every day, while other people can lead reclusive lives until the pandemic is over. Our models (5-7) and (19) do not feature such structuring. The process for incorporating structuring has been demonstrated in Ref. [17], and we leave the consideration of the results for future study.

### §3 ADDITIONAL RESULTS AND DISCUSSION

The Article proper has space for only the most impactful results; here we investigate different parameter combinations, present the epidemic time traces and discuss several other issues which we think are of strategic interest.

#### NO TRAVEL

Table 1 of the Article proper contains a paradoxical observation. In eight of the nine situations considered, the epidemic duration is under 400 days, but in the worst case (least effective vaccine, most transmissible virus) it is almost 1000 days. How did this happen ? To resolve this, we plot the epidemic time trace in this instance, as Figure S1 below.

**Figure S1:**
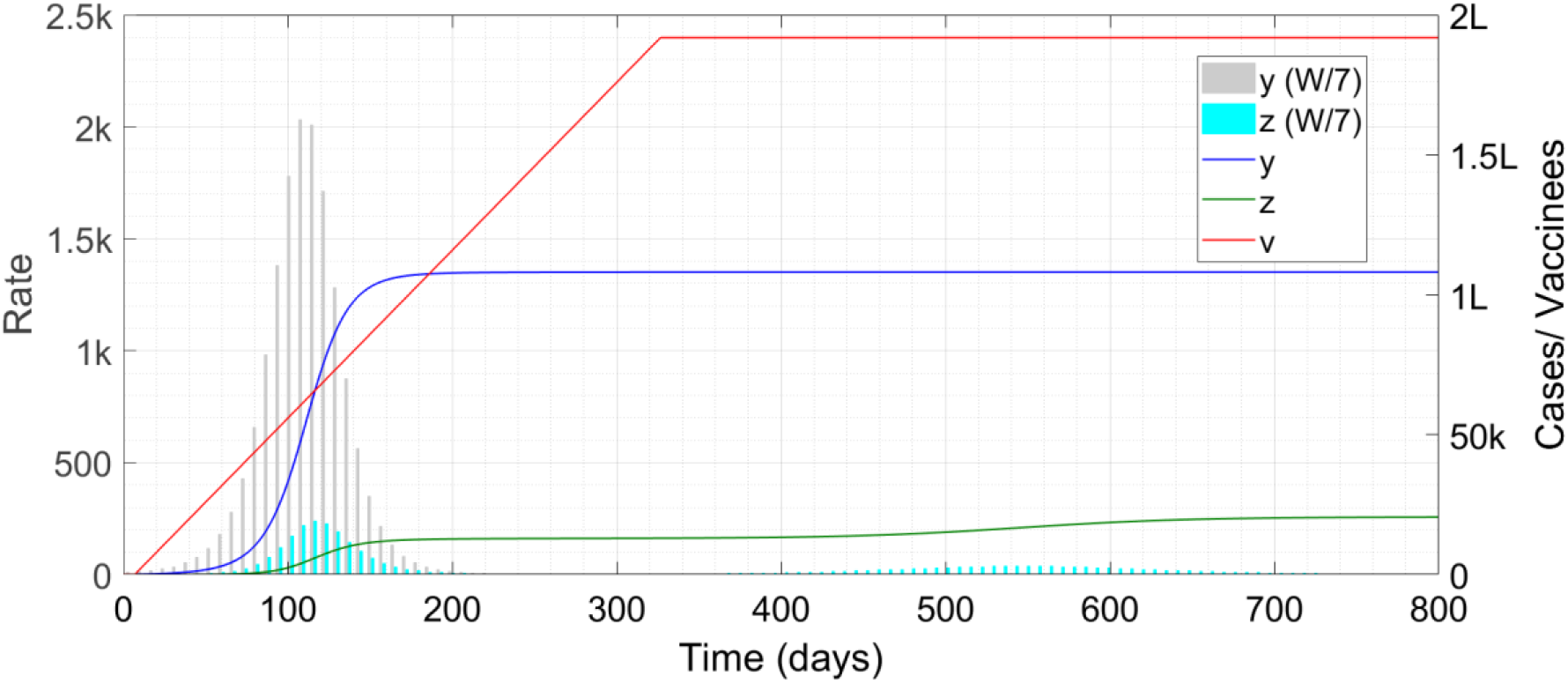
Time trace of evolution of the epidemic for the worst-case situation considered in Table 1 i.e. η = 60 percent and vaccinee mobility R_eq_ = 5. Blue line denotes the unvaccinated cases, green line the vaccinated cases and red line the total number of vaccinees. Grey and cyan bars denote the epidemiological curve or epi-curve, i.e. the weekly case counts in the unvaccinated and vaccinated groups respectively. We have scaled them down by a factor of seven so that the envelope of the bars might be the derivatives 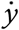 and ż. The legend W/7 indicates this. ‘k’ denotes thousand and ‘L’ hundred thousand.

We can see that there is a second wave of cases in the vaccinated group, starting about the time the vaccination drive stops. With partially effective vaccine, a vaccination drive which clears people to higher mobility acts like a gradual reopening instead of a control measure, and fuels a second wave. Reopening-induced second waves have been characterized in our prior work [20] and have been seen in USA and many countries in western Europe. Here we see a similar phenomenon, which explains the anomalously high epidemic duration.

As discussed in §1, we have characterized public health interventions through the spreading rate *m*, which itself is a product of two numbers – the interaction rate and the transmission probability conditional on susceptible target. In writing the equations (5,6), we have assumed that the interaction rate is low for non-vaccinees and high for vaccinees, that these interactions are distributed among the two groups in proportion to their populations and that the transmission probability given target susceptibility is constant independent of the nature of the target. These assumptions were motivated partly by a desire for simplicity; we could also have written (5,6) as follows :

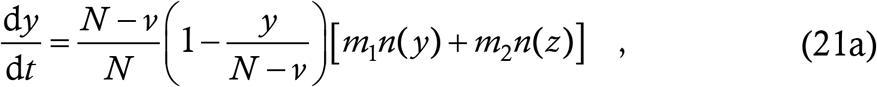

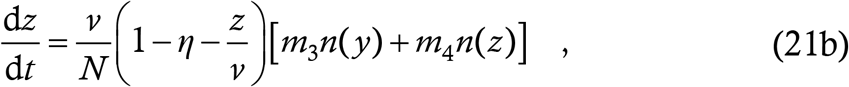

which features four *m*’s instead of two.

Equation (21) can account for situations like vaccinees preferring to interact only with other vaccinees, wearing masks in the presence of non-vaccinees etc. In an ideal situation, *m*_1_, *m*_2_ and *m*_3_ all ought to equal *m*_*l*_ and only *m*_4_ equal *m*_*h*_ – vaccinees should in principle observe all restrictions such as minimizing interactions and masking whenever there are non-vaccinees around. This situation would lead to lower case counts than what we have found in Table 1 of the Article proper (which has *m*_1_ = *m*_3_ = *m*_*l*_ and *m*_2_ = *m*_4_ = *m*_*h*_). In a worst-case situation however, only *m*_1_ may remain *m*_*l*_ and all of *m*_2_, *m*_3_ and *m*_4_ become *m*_*h*_ – this will happen if neither vaccinees nor non-vaccinees feel any qualms about interacting freely with the other class, believing that one vaccine will take care of two people. This would lead to higher case counts than our estimates.

To analyse this in more detail, we consider the situations of Table 1 in the above best- and worst-case scenarios. That is, we take the same *m*_*l*_ (0·186 corresponding to *R*_*eq*_=1·15), the same three vaccine efficacies of 60, 75 and 90 percent and the same three values of *m*_*h*_ (*R*_*eq*_’s of 2, 3 and 5), and calculate the durations and case counts when (*a*) “B” : *m*_1_ = *m*_2_ = *m*_3_ = *m*_*l*_, *m*_4_ = *m*_*h*_ and (*b*) “W” : *m*_1_ = *m*_*l*_, *m*_2_ = *m*_3_ = *m*_4_ = *m*_*h*_. We present the results in Table S1.

**Table S1:**
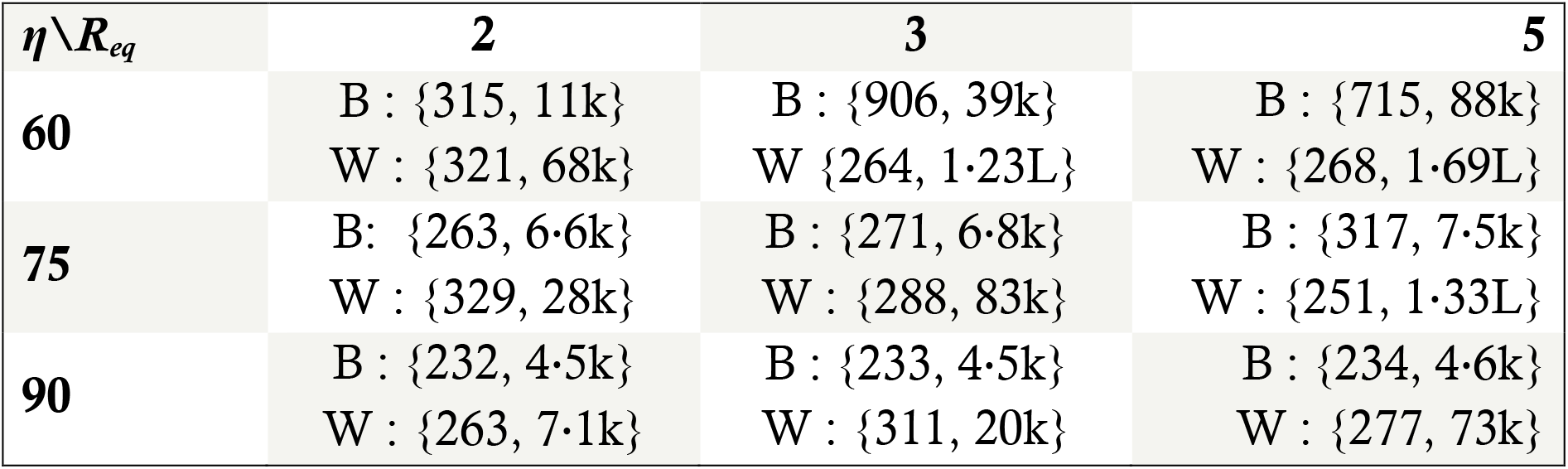
Best (B) and worst (W) case scenarios with respect to vaccinee and non-vaccinee mobility for the situations considered in Table 1 of the Article proper. For each scenario we show the epidemic duration T (days) and the cumulative case count X in curly brackets, thus {T, X}. The symbol ‘k’ denotes thousand and ‘L’ hundred thousand.

For 75 and 90 percent vaccine efficacy, we can see a significant difference between the infection counts in the best- and worst-case scenarios, with the values given in Table 1 of the main Article lying in between these two extremes, which is consistent. In fact, for *R*_*eq*_ = 5, the gap between the various outcomes is tremendous. It shows very strong rewards to be had if vaccinees relax their guard only in the presence of other vaccinees and not in the presence of non-vaccinees. We can also see however that this difference is absent with the 60 percent effective vaccine – only the best-case scenario with the lowest *R*_*eq*_ features an acceptable caseload while all the other situations involve high caseload.

If 75 makes a good vaccine and 60 a bad vaccine, then this leads to another question – what is the cutoff ? How does the infection control performance depend on vaccine efficacy ? In Tables 1 and S1, we had space for only three discrete efficacies; what happens if we were to treat the efficacy as a continuous variable ? To gain insight, we consider the interaction situation of Table 1 (not S1), fix vaccinee *R*_*eq*_ = 3 and plot the duration *T* together with the total case count *X* as a function of vaccine efficacy between 50 and 100 percent. This plot is shown in Figure S2 below.

**Figure S2:**
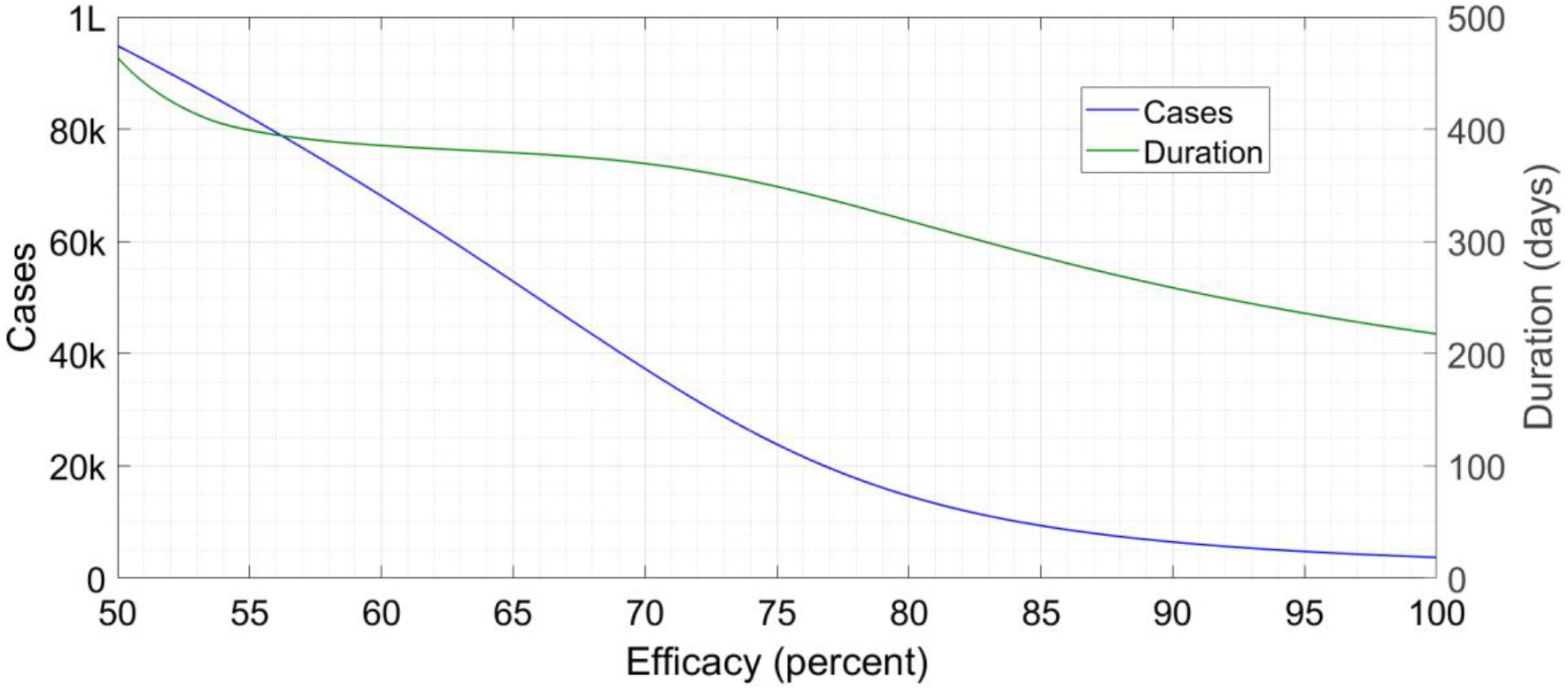
Cumulative caseload (blue) and epidemic duration (green) as functions of the vaccine efficacy.

We can see that as the efficacy decreases from 100, the caseload features a slow increase upto about 85 percent efficacy, followed by a much sharper increase as efficacy reduces below 75. Thus, for this parameter combination, 80 or 85 percent may be treated as a “cutoff” for acceptable efficacy. This cutoff will vary depending on the interaction parameters – for example if we replot Figure S2 with vaccinee *R*_*eq*_ = 5 and the best-case interaction situation from Table S1, then we get the following (Figure S3).

**Figure S3:**
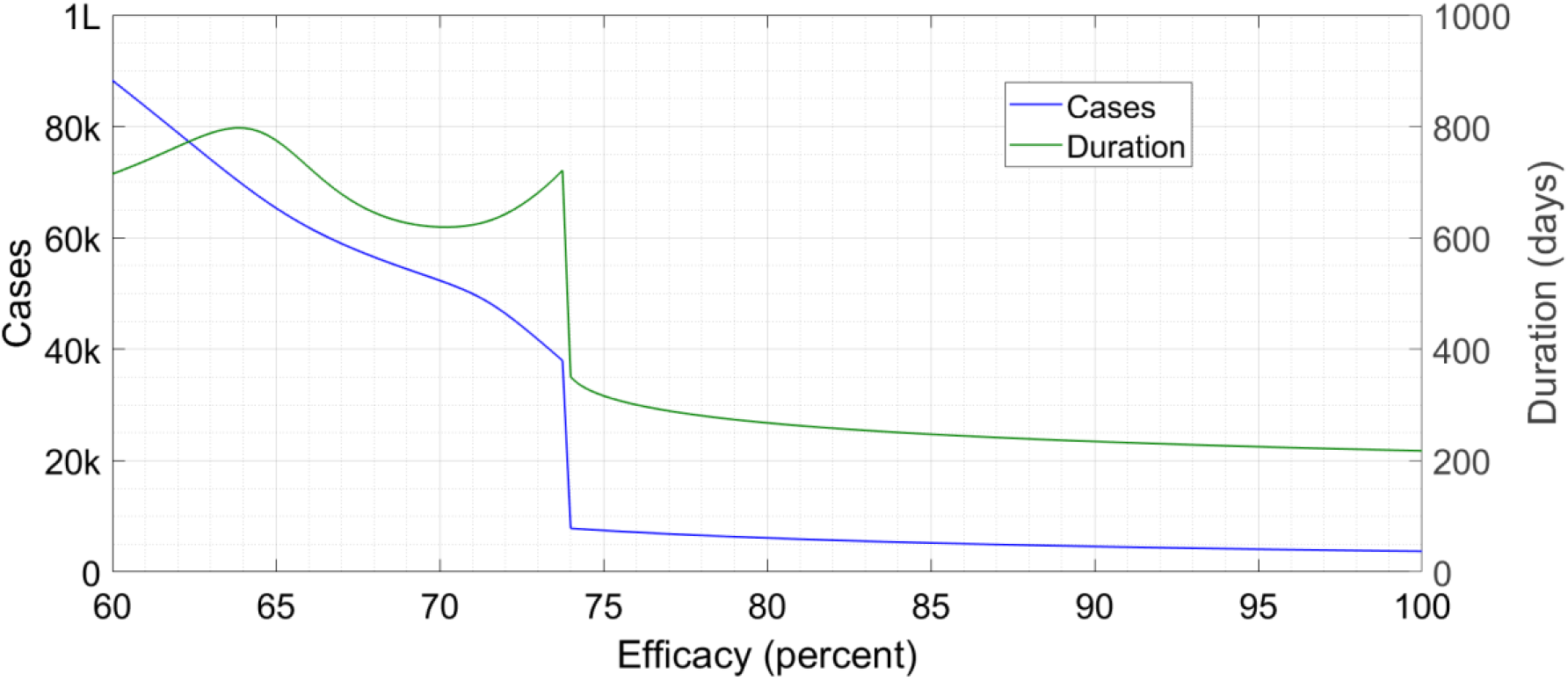
Cumulative caseload (blue) and epidemic duration (green) as functions of the vaccine efficacy.

In this case, we can see a big jump in the curves at 74 percent vaccine efficacy – this jump corresponds to a bifurcation where a one-shot solution (Figure 1) cedes to a two-wave solution (Figure S1). Before the jump i.e. at high efficacy, caseload is almost independent of efficacy while after the jump it increases rapidly with decreasing efficacy. Evidently, with this interaction situation, 74 is the cutoff efficacy – note that the argument based on *R* in the Discussion Section of the Article proper would have predicted this cutoff to be 80.

There are two ways of hastening the end of the epidemic beyond the durations we are finding here. One is to increase the rate of vaccination. For example, doubling the vaccination rate for the situation of Figure 1 in the Article proper causes the epidemic to end at 264 days instead of 352. We hope that as time elapses, more and more vaccines become available so that the vaccination drive and hence the elimination efforts can increase in pace. The second way is preferential vaccination of shopkeepers, bankers, bus conductors etc who are forced to interact with many people as part of their jobs. This strategy has been mentioned in several recent studies [7-10] and can lead to substantial gains in time as well as caseload.

We note that with selective relaxation, the end of the epidemic is being brought about not by achieving high vaccination coverage (herd immunity) but by slowly and systematically starving the virus of new targets in both the unvaccinated and vaccinated groups. To see this, consider the case *η* = 90 percent and vaccinee *R*_*eq*_ = 3 from Table 1. The outbreak ends at 259 days by which time 1,55,000 vaccines have been distributed. The total number of immune people (successfully immunized vaccinees plus actual cases) is 1,46,000 which is significantly less than the 67 percent immunization level required to achieve herd immunity with an *R* of 3. As another example, with *η* = 75 percent and vaccinee *R*_*eq*_ = 5 in Table S1, the immunized fraction at the end is almost exactly 50 percent and not 80 percent.

#### TRAVEL

We first estimate typical values of the amplification factor *k* for normal (mask-free) travel situations. For air travel, each window/aisle seater case will infect the middle seater, while each middle seater case will infect both the window and aisle. Even though the ventilation in an aircraft is good [S4], normal travel involves conversation and occasional physical contact between neighbouring passengers. Furthermore, each infected passenger will likely infect two others (nearest neighbours) in the airport seating area. There might be two further neighbours in the boarding queue but the exposure time here will be small and transmission will be less than certain in this case. Overall, very approximately, air travel might generate a *k* in the range 4-6. We can expect an analogous situation for rail travel in western countries, where the seating is similar. In Indian Railways, the world’s busiest railway network, the AC First class features four people in a compartment, the AC 2-Tier class features six and the AC 3-Tier and Sleeper classes feature eight. In these situations we can expect approximate *k* values of 3, 5 and 7 respectively.

We now present a few time traces of the epidemic evolution for the situations considered in Tables 2 and 3 of the Article proper. First, we take an instance where City 2 has negligible impact on City 1. Consider the *γ* = 20 and *k* = 5 situation from Table 2. In Figure S4, we show the time traces of the two cities with no travel.

**Figure S4:**
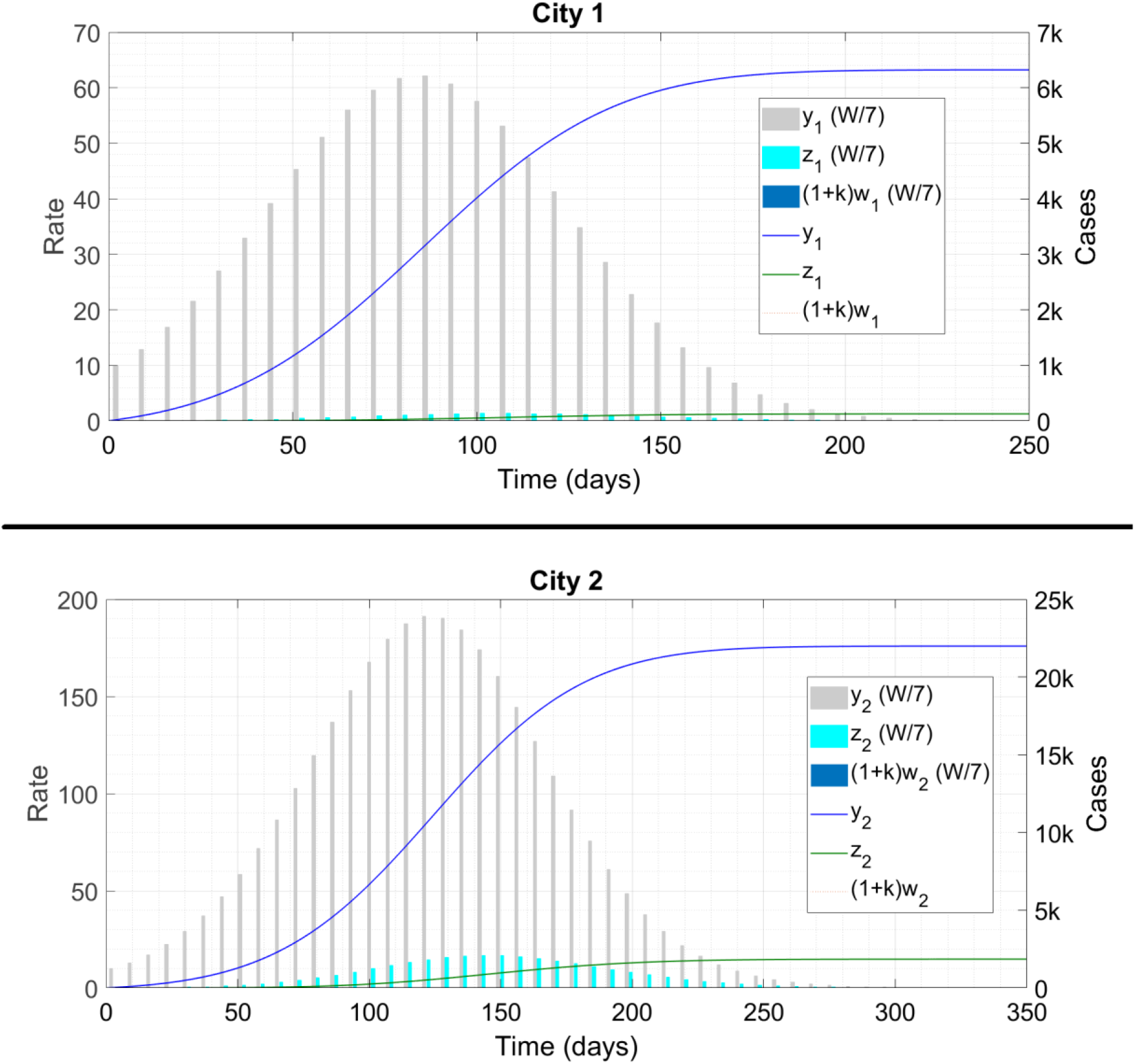
Time traces of the epidemic in City 1 (vaccinee R_eq_ = 3 and η = 90 percent) and City 2 (R_eq_ = 3 and η = 75 percent) when there is no travel. In each plot, blue line denotes unvaccinated cases, while green line denotes vaccinated cases; the corresponding epi-curves are in grey and cyan respectively. The quantities w_1_ and w_2_ appear in the plot legend but not in the plot itself because they are both zero, as they should be. ‘k’ denotes thousand and W/7 weekly cases scaled down by a factor of seven.

The time trace of City 1 here is the same as Figure 1 of the main Article, which acts as a check on our numerical work. The primary difference between the two cities lies in the time when they attain their respective peaks – cases in City 1 peak at about 80 days while vaccine cases in City 2 (the ones who travel) peak at about 150 days. With travel coupling, the significant quantity is the spreading profile in City 1, so we present that below as Figure S5.

**Figure S5:**
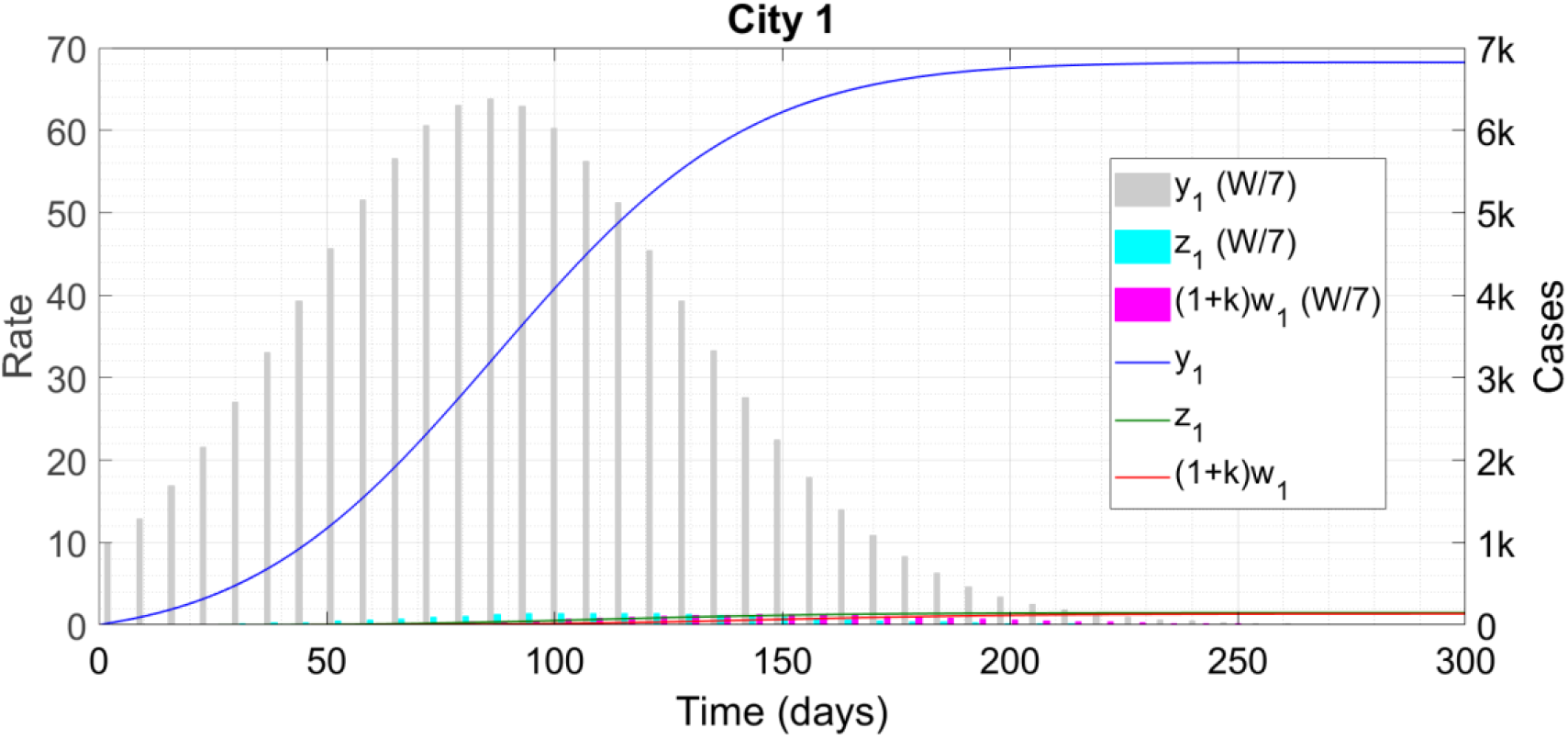
Time traces of the epidemic in City 1 (R_eq_ = 3 and η = 90 percent) when there is travel to and from City 2. Blue line denotes unvaccinated cases, green line vaccinated cases and red line travelled cases – for the last one, we club together the imported and vehicle-generated cases. The corresponding epi-curves are in grey, cyan and magenta respectively. ‘k’ denotes thousand and ‘W/7’ weekly cases scaled down by a factor of seven.

We can see that the peak in City 1 is also not delayed on account of the influx.

For another example, we take an instance where City 2 completely spoils City 1’s infection control performance. Consider Table 3, where City 1 remains as it was, but the vaccine efficacy in City 2 is reduced to 60 percent. In particular, focus on the ‘worst’ cell where *γ* = 100 and *k* = 10. In Figure S6 we present a plot of City 2’s performance in the absence of travel.

**Figure S6:**
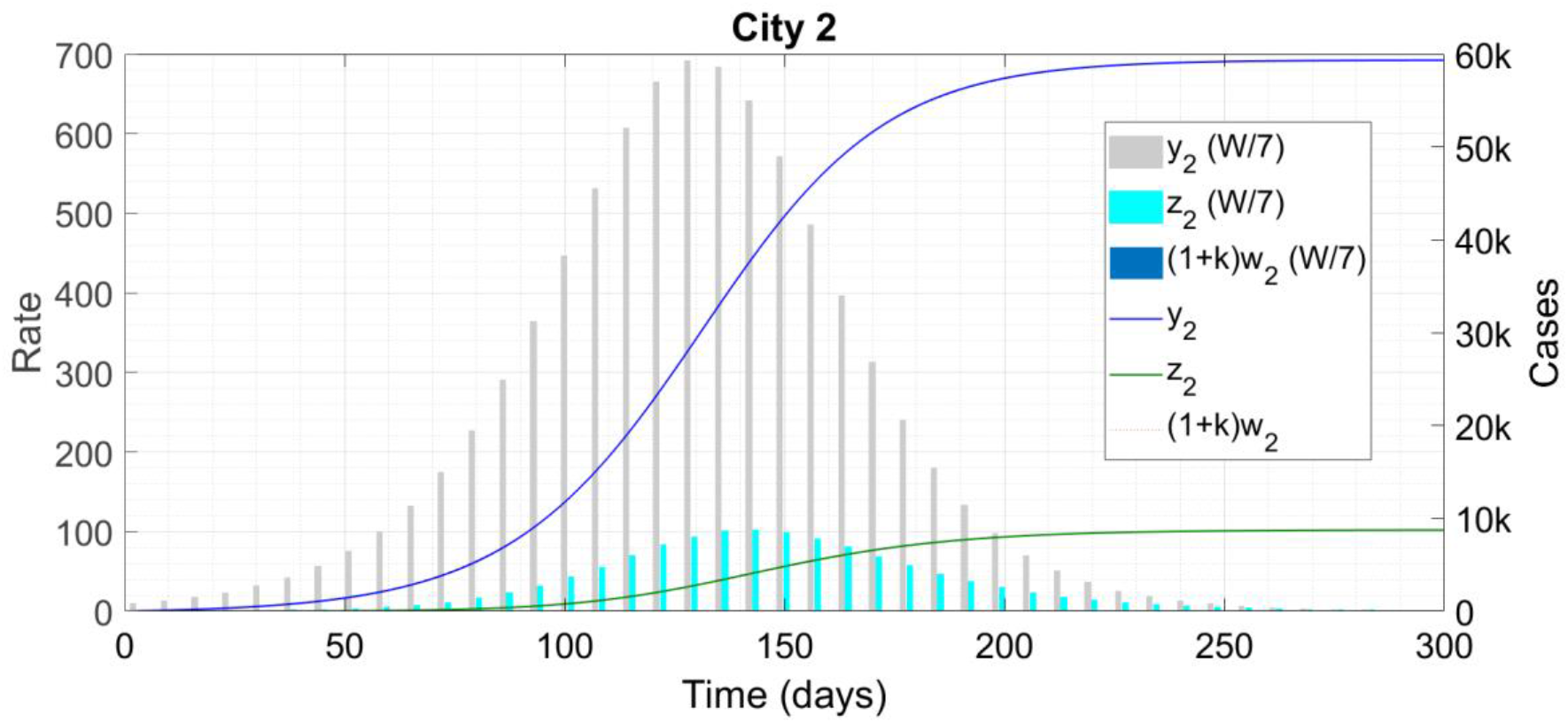
Time traces of the epidemic in City 2 (vaccinee R_eq_ = 3 and η = 60 percent) when there is no travel. Blue line denotes unvaccinated cases and green line vaccinated cases. The corresponding epi-curves are in grey and cyan respectively. The quantities w_1_ and w_2_ appear in the plot legend but not in the plot itself because they are both zero. ‘k’ denotes thousand and ‘W/7’ weekly cases scaled down by a factor of seven.

This is qualitatively similar to Figure S4 (bottom panel) except that the actual numbers involved are much higher. Now we present the trajectories in City 1 when coupled strongly to this rogue city (Figure S7).

**Figure S7:**
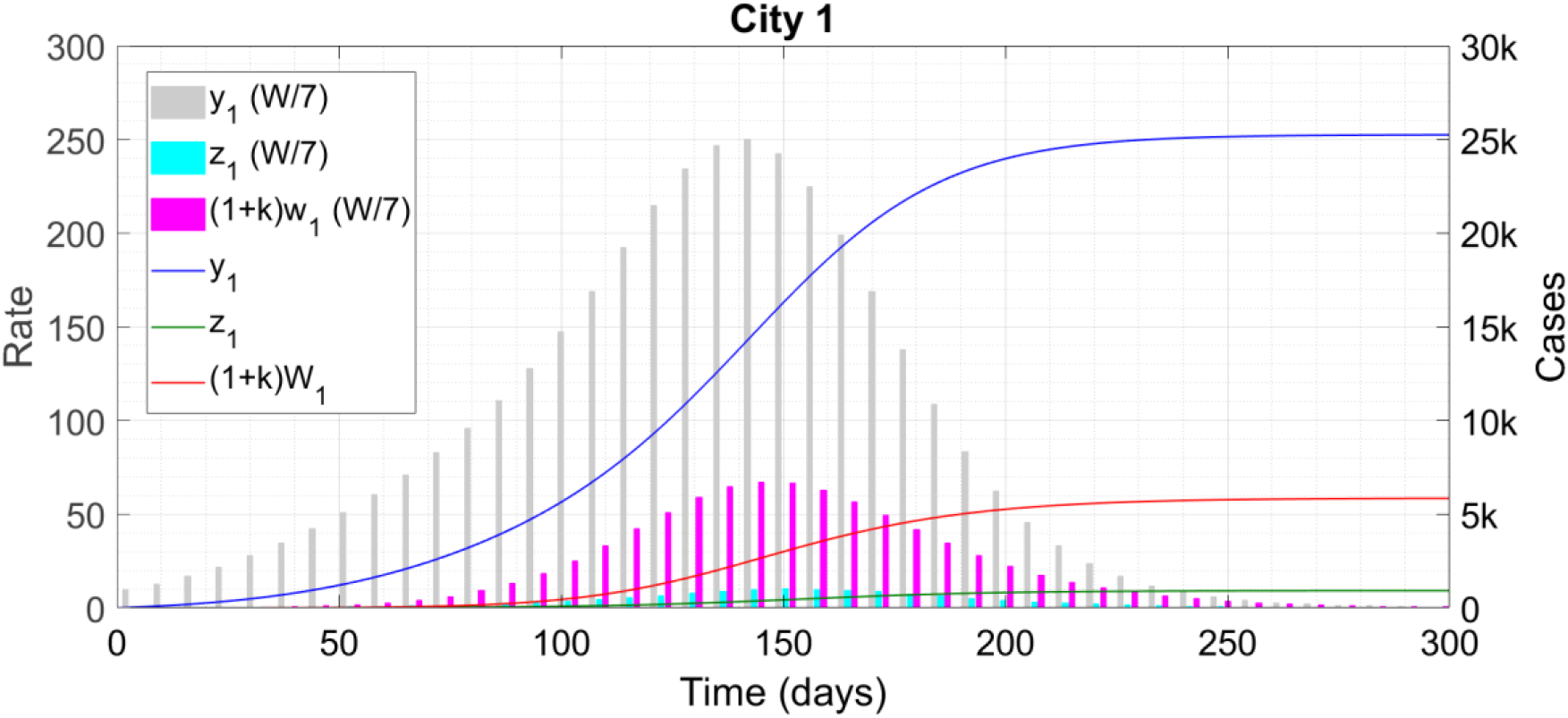
Time traces of the epidemic in City 1 (R_eq_ = 3 and η = 90 percent) when there is travel to and from City 2 (R_eq_ = 3 and η = 60 percent). Blue line denotes unvaccinated cases, green line vaccinated cases and red line travelled cases – for the last one, we club together the imported and vehicle-generated cases. The corresponding epi-curves are in grey, cyan and magenta respectively. ‘k’ denotes thousand and ‘W/7’ weekly cases scaled down by a factor of seven.

Here we can clearly see the effect of the travel. City 1 starts off as it did in the absence of travel (Figure S4) but by about the 100-day mark, travel is introducing as many new cases as it was generating on its own. So, instead of flattening out, the curve now becomes enslaved to that of City 2’s vaccinees and peaks together with City 2. Only when the disease comes under control in City 2 does it also follow suit in City 1.

### SOCIOECONOMIC AND POLICY ASPECTS

With an effective vaccine, immediate and preferential relaxation of restrictions for vaccinees appears to be a quick and surefire path to elimination of COVID-19 in time while achieving maximum socioeconomic recovery. The selective relaxations however may cause negative emotions between people who get vaccinated earlier and those who get vaccinated later. To the largest extent, there is nothing to be done about this rift – while vaccine allocation policies can come up with a priority order which maximizes the common good, it will be inevitable that one healthy young well-paid software engineer will get the shots and hence a ticket to freedom two months before another healthy young well-paid software engineer.

The negativity arising from such heterogeneity can be alleviated through a public information campaign – the impact of the virus itself is very heterogeneous and that is outside our control, so a bit of manmade inequality during the endgame phase is also tolerable. This is especially true since continuing blanket restrictions until the disease has been eliminated will entail tremendous financial losses on the part of the travel industry and many other businesses. Nonetheless, to avoid unhealthy competition, employers and universities who arrange for vaccination of employees and/or students might wait to initiate the general vaccination drive (excluding frontline, high-interaction and high-risk people) until they have received all the requisite doses.

During selective relaxation, public health authorities will have to work to ensure that minimal close and unmasked interaction occurs between vaccinees and non-vaccinees – as we saw by comparing Tables 1 and S1, a lot of gain can be achieved through this. To facilitate this, vaccinees may be given apparel or badges which prominently advertise their status. In situations where physical segregation of the two classes is impossible, like shops and restaurants, mask and separation requirements might need to remain in place for the vaccinees as well. If a chain store or eatery has multiple similar outlets in the same city, like McDonald’s, it might designate some as vaccinee only with no restrictions and others as common spaces with restrictions. In airports, regions occupied by vaccinees should be screened off from those occupied by non-vaccinees. In places where this is not possible, for example at the security checking area (and it will almost certainly be impossible on railway platforms), vaccinees should also observe all precautions. Masking at just the frisking queue or the station is a negligible inconvenience compared with masking and face-shielding throughout a flight or a long-distance train.

A plethora of results supports the assertion that a vaccine of 80 percent or higher efficacy can act as the basis for an immunity passport while a vaccine of 60 percent efficacy cannot, unless the virus transmission rate is already low. Hence, research into development of more efficacious vaccines should continue even as the early candidates are administered. The efficacies of existing vaccines should also be monitored for socio-demographic deter-minants if any. For example, if Vaccine A has 90 percent efficacy in the 18-50 age group and 40 percent in the 65+ age group while Vaccine B has 70 percent across all age groups, then younger people should be given Vaccine A and older people Vaccine B. One way of measuring efficacy “in the field” as opposed to in a placebo-controlled trial setting is by using the following definition, which yields the efficacy *E*(*t*) as a function of time :

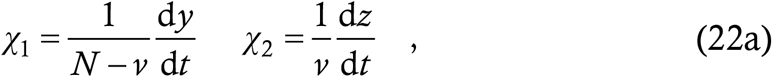

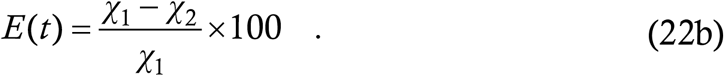

It has been shown in Ref. [S5] that this definition yields *E*(*t*) very close to *η* during the bulk evolution of the epidemic.

A placebo-free definition of efficacy can also help with expediting vaccine trials. Phase 3 trials are an essential component for determining efficacy, and those require the voluntary participation of 25,000 people or more. As approved vaccines become increasingly available, people will be correspondingly reluctant to participate in a trial for a new candidate, even if it promises better results. To encourage participation, the placebo arm of the trials may be deferred and all trial participants offered access to the vaccine, so that they can at least have “jumped the queue” if the undertrial vaccine does secure approval. Equation (22) may be used to compute the efficacy from such a trial.

In conclusion, we look forward to the day when we can get an effective vaccine, doff our mask and hop onto a flight headed back to life as we knew it, and we hope that this happy day arrives sooner rather than later.

